# Older age is associated with decreased sensitivity to stress and severity of anxiety, depression, and post-traumatic stress disorder: Evidence from the Ukrainian population

**DOI:** 10.1101/2025.07.29.25332353

**Authors:** Oleh Lushchak, Svitlana Bolman, Pavlo Petakh, Oleksandr Kamyshnyi, Olha Strilbytska

## Abstract

Sustained exposure to violence, displacement, and chronic uncertainty caused by the ongoing Russian aggressive war affected millions of civilians, prompting serious concerns regarding the cumulative and long-term effects on the mental health of the population in wartime conditions. This study aims to assess the prevalence of stress, depression, anxiety, and post-traumatic stress disorder (PTSD) symptoms among the Ukrainian population after two years of Russian invasion. In addition, the study examines the association between age and the level of psychological symptoms, aiming to identify age-specific trajectories in the development and expression of war-related stress responses. Using a cross-sectional survey design, data were collected from a diverse sample of Ukrainian participants across different age groups. The study included a total of 9,967 Ukrainian participants, devided into five age groups: 1,226 individuals (12.3%) aged 18-24 years, 4,016 (40.3%) were aged 25-34 years, 3,227 individuals (32.4%) were aged 35-44; 1,171 respondents (11.8%) were between 45 and 54 years old; and 327 participants (3.3%) were 55 years old and above. Standardised psychometric tools were used to assess the severity of psychological symptoms across participants. Stress levels were measured using the Perceived Stress Scale (PSS-10), anxiety symptoms were evaluated with the Generalised Anxiety Disorder scale (GAD-7), depression levels were estimated with the Patient Health Questionnaire (PHQ-9) and post-traumatic stress disorder symptoms were assessed using the PTSD Checklist for DSM-5 (PCL-5). The results reveal that stress levels tend to be lower in older adults, suggesting that older individuals may possess greater psychological resilience or employ more adaptive coping strategies. In contrast, younger respondents were found to exhibit significantly higher levels of anxiety, depression and PTSD symptoms, indicating a heightened vulnerability to the traumatic effects of war among youth. Analysis of differential symptom profiles across age cohorts will further help to uncover potential age-linked risk and resilience factors that modulate the psychological impact of prolonged armed conflict.

---

Post-traumatic stress disorder (PTSD) is a mental health condition that can arise after a person experiences or witnesses highly distressing or life-threatening events, including warfare, violence, serious accidents, or natural disasters. PTSD is a commonly studied condition in populations exposed to war, conflicts, and displacements (Steel et al., 2009). Indeed, depression and PTSD have a point prevalence rate of 26.51% among adult survivors who stay in war-afflicted regions (Morina et al., 2018). These concerns take on heightened relevance in the context of Ukraine, where the civilian population has been exposed to prolonged and large-scale traumatic experiences following Russia’s full-scale invasion in February 2022. However, even before the 2022 conflict started, Ukrainians were already dealing with PTSD (Shevlin et al., 2018). One in five (22%) people who have experienced war or other conflicts in the previous 10 years will have depression, anxiety, post-traumatic stress disorder, bipolar disorder, or schizophrenia (WHO response to the Ukraine crisis, 2023). In applying these estimates to Ukraine, the World Health Organisation (WHO) expects that approximately 9.6 million people in Ukraine may have a mental health condition (WHO response to the Ukraine crisis, 2023).

Several risk factors, including female gender, older age, and uncoupled residing, have been proven to be associated with psychological distress (Viertiö et al., 2021). However, it was generally reported in epidemiological studies that the lower prevalence of PTSD is observed in older adults relative to younger adults. Indeed, PTSD is most common among individuals between the ages of 45 and 59 years old (9.2%) as compared to individuals above the age of 60 years old (2.8%) in the United States (National Comorbidity Survey, 2007, Harvard Medical School). National Epidemiologic Survey on Alcohol and Related Conditions (NESARC-2) showed that older adults are more likely to have traumatic experiences than middle-aged and younger adults (Reynolds et al., 2016). Similar patterns of lower prevalence of PTSD and trauma exposure among older compared with younger adults have been reported in several countries (Böttche et al., 2011). Moreover, men and women differed in the age-specific distribution of PTSD (Ditlevsen and Elklit, 2010). The prevalence of PTSD was highest, approximately in the 40s for men and in the 50s for women (Ditlevsen and Elklit, 2010). However, the study by Kessler and colleagues (1999) showed that the prevalence of PTSD was highest from the mid-40s to mid-50s in males and from the mid-20s to mid-30s in females.

The prevalence of PTSD is significantly higher in the military as compared to civilian populations. PTSD is more common among U.S. military veterans between the ages of 18 and 29 years at 29.3%, compared to veterans over the age of 60 years at 4% (Wisco et al., 2016).

While there is a strong confirmation of low PTSD prevalence in older people (Frueh et al., 2007), some studies contradict previous research (Maercker et al., 2009). The study by Maercker and colleagues (2009) showed a linear increase in the prevalence of PTSD during aging in the German population, which is assumed to be related to the consequences of World War II. The participants in the age range of 60 to 93 years displayed significantly higher PTSD prevalence as compared to participants younger than 60 years (Maercker et al., 2009). Traumatic events and daily stress also have a psychological impact on adolescents living in a war-torn region. Some studies have shown that older children reported higher levels of distress than younger children (El-Khodary et al., 2020). However, other data claimed opposite effects (Allwood et al., 2002). Variations of periods of military service, the nature of trauma exposure and the sociohistorical context surrounding a war might have contributed to the inconsistent findings about PTSD prevalence. Moreover, wide variations could be attributed to methodological issues and different psychiatric assessment instruments.

To our knowledge, there are several empirical psychological studies of the civilian population conducted during the course of the hostilities. The study by Karatzias and colleagues (2023) did not find age differences in war-related experiences. The author suggested that the restricted age profile of the sample may be a limitation of this study. PTSD prevalence is the lowest for younger adults and the highest for individuals over 40 (Zasiekina et al., 2023). However, some studies suggested that PTSD prevalence depends mostly on social, economic, and cultural context, and personal characteristics rather than age (Ozer et al., 2003). Further research is needed in this area to establish if there are associations between PTSD symptoms and age during war.

This study examines the impact of the Russia-Ukraine war on the mental health of various age cohorts of Ukrainian civilians who live under continuous traumatic stress. Conducted during the second year of the full-scale invasion, it captures the mental health effects of prolonged traumatic stress, focusing specifically on age as a differentiating factor. Our previous nationally representative study, which examined PTSD prevalence across displacement status – non-displaced persons (NDPs), internally displaced persons (IDPs), and refugees abroad (Lushchak et al., 2023). The present study explores the mental health burden across five distinct age cohorts within the population of Ukraine. They also aimed to establish whether age as a sociodemographic factor is associated with positive scores for stress, anxiety, depression, and PTSD among Ukrainians during the ongoing war.

## RESULTS

### Survey Sample Characteristics

A total of 11,014 individuals of Ukrainian origin participated in the survey. However, responses from 1,047 individuals were excluded because they were under 18 years of age or residing outside Ukraine at the onset of the Russian invasion in 2022. Further data quality checks included screening for patterned or “placeholder” responses, as well as response styles indicative of extreme or midpoint bias. The final analytical sample consisted of 9,967 adult respondents, aged between 18 and 81 years.

Participants were grouped according to their age at the time of the survey. Specifically, 1,226 individuals (12.3%) were aged 18-24 years; 4,016 participants (40.3%) fell into the 25-34 age group; 3,227 individuals (32.4%) were aged 35-44; 1,171 respondents (11.8%) were between 45 and 54 years old; and 327 participants (3.3%) were aged 55 and above. Demographically, females constituted the majority across all groups, with a particularly high representation among participants of the 35-44 age group (92.2%).

As expected, the level of education varies by age. The highest proportion of participants with secondary school education is observed in the 18-25 age group (19.3%), while the lowest is found among those aged 55 and above (1.2%). The proportion of individuals holding an academic degree increases with age, reaching up to 18% among participants aged 55 and older. Age was found to be significantly associated with both the likelihood and number of children. The analysis reveals a clear age-related trend, with the proportion of individuals reporting at least one child increasing from 2.8% among those aged 18-24 to 44.6% among those aged 55 and older.

The highest proportion of non-displaced individuals in the context of war-related migration was found among the population aged 55 years and older (73.7%).

### Overview of Mental Health Status

Table 2 summarises the prevalence of previously diagnosed mental health conditions among respondents across five age categories. Diagnoses were self-reported and reflected conditions identified before the survey. The data show that the highest level of receiving a mental health diagnosis at some point in the past is among participants of the 18-24 age group (69.6%). At the same time, the lowest prevalence of mental health issues was recorded among individuals aged 55 and above (41.9%).

**Table 1:** Sociodemographic Characteristics and Group Comparison.

|  | **18-24**<br>**n = 1226**<br>**(12.3%)** | **25-34**<br>**n = 4016**<br>**(40.3%)** | **35-44**<br>**n = 3227**<br>**(32.4%)** | **45-54**<br>**n = 1171**<br>**(11.7%)** | **55+**<br>**n = 327**<br>**(3.3%)** | **Cramer's**<br>**V** | **p value** |
| --- | --- | --- | --- | --- | --- | --- | --- |
| **Sex** |  |  |  |  |  |  |  |
| Female | 1014<br>(82.7%) | 3632 (90.4%) | 2976<br>(92.2%) | 1057<br>(90.3%) | 284 (86.9%) | 0.0971 | <.001 |
| Male | 212 (17.3%) | 384 (9.6%) | 251 (7.8%) | 114 (9.7%) | 43 (13.1%) |  |  |
| **Marital status** |  |  |  |  |  |  |  |
| Not married | 986 (80.4%) | 1513 (37.7%) | 575 (17.8%) | 131 (11.2%) | 28 (8.6%) | 0.268 | <.001 |
| Married | 172 (14.0%) | 2211 (55.1%) | 2222<br>(68.9%) | 787 (67.2%) | 181 (55.4%) |  |  |
| Divorced | 6 (0.5%) | 171 (4.3%) | 358 (11.1%) | 221 (18.9%) | 79 (24.2%) |  |  |
| Other | 62 (5.1%) | 121 (3.0%) | 72 (2.2%) | 32 (2.7%) | 39 (11.8%) |  |  |
| **Kids** |  |  |  |  |  |  |  |
| No Children | 1188<br>(97.0%) | 2711 (67.5%) | 954 (29.6%) | 159 (13.6%) | 23 (7.0%) | 0.546 | <.001 |
| 1 Child | 34 (2.8%) | 992 (24.7%) | 1190<br>(36.9%) | 472 (40.3%) | 146 (44.6%) |  |  |
| 2 Children | 2 (0.2%) | 276 (6.9%) | 882 (27.3%) | 444 (37.9%) | 141 (43.2%) |  |  |
| 3+ Children | 0 (0.0%) | 37 (0.9%) | 201 (6.2%) | 96 (8.2%) | 17 (5.2%) |  |  |
| **Education level** |  |  |  |  |  |  |  |
| Secondary school | 237 (19.3%) | 88 (2.2%) | 55 (1.7%) | 16 (1.4%) | 4 (1.2%) | 0.206 | <.001 |
| Secondary specialized education | 118 (9.6%) | 245 (6.1%) | 197 (6.1%) | 140 (12.0%) | 32 (9.8%) |  |  |
| Higher education | 871 (71%) | 3557 (88.6%) | 2708<br>(83.9%) | 861 (73.5%) | 232 (70.9%) |  |  |
| Scientific degree | 0 (0.0%) | 126 (3.1%) | 267 (8.3%) | 154 (13.2%) | 59 (18.0%) |  |  |
| **Have been in zone of active hostilities** |  |  |  |  |  |  |  |
| As a civilian | 241 (19.7%) | 969 (24.1%) | 863 (26.7%) | 279 (23.8%) | 69 (21.1%) | 0.0398 | <.001 |
| As a military | 5 (0.4%) | 32 (1.4%) | 16 (0.5%) | 9 (0.8%) | 1 (0.3%) |  |  |
| None | 980 (79.9%) | 3015 (75.1%) | 2348(72.8%) | 883 (75.4%) | 257 (78.6%) |  |  |
| **Status** |  |  |  |  |  |  |  |
| Refugees | 168 (13.7%) | 727 (18.1%) | 715 (22.2%) | 187 (16.0%) | 35 (10.7%) | 0.0777 | <.001 |
| IDPs | 89 (7.3%) | 414 (10.3%) | 258 (8%) | 83 (7.1%) | 29 (8.9%) |  |  |
| NDPs | 827 (67.5%) | 2342 (58.3%) | 1699<br>(52.6%) | 768 (65.6%) | 241 (73.7%) |  |  |
| Returnees | 142 (11.6%) | 533 (13.3%) | 555 (17.2%) | 133 (11.4%) | 22 (6.7%) |  |  |

**Table 2:** Mental Health Status of Various Age Groups.

|  | **18-24**<br>**n = 1226**<br>**(12.3%)** | **25-34**<br>**n = 4016**<br>**(40.3%)** | **35-44**<br>**n = 3227**<br>**(32.4%)** | **45-54**<br>**n = 1171**<br>**(11.7%)** | **55+**<br>**n = 327**<br>**(3.3%)** | **Cramer's**<br>**V** | **p value** |
| --- | --- | --- | --- | --- | --- | --- | --- |
| *Previously diagnosed mental health issues* |  |  |  |  |  |  |  |
| **Anxiety** | 359 (29.3%) | 1139 (28.4%) | 757 (23.4%) | 232 (19.8%) | 58 (17.7%) | 0.0801 | <.001 |
| **Depression** | 274 (22.4%) | 959 (23.9%) | 596 (18.5%) | 188 (16.1%) | 49 (15.0%) | 0.0765 | <.001 |
| **PTSD** | 90 (7.3%) | 308 (7.7%) | 218 (6.8%) | 72 (6.1%) | 18 (5.5%) | 0.0236 | 0.236 |
| **Other** | 130 (10.6%) | 279 (6.9%) | 145 (4.5%) | 41 (3.5%) | 12 (3.7%) | 0.0898 | <.001 |
| **None** | 754 (61.5%) | 2471 (61.5%) | 2188 (67.8%) | 843 (72%) | 245 (74.9%) | 0.0886 | <.001 |
| *Received mental health care* |  |  |  |  |  |  |  |
| **Regularly** | 212 (17.3%) | 937 (23.3%) | 600 (18.6%) | 115 (9.8%) | 11 (3.4%) | 0.0987 | <.001 |
| **Sometimes** | 267 (21.8%) | 910 (22.7%) | 785 (24.3%) | 262 (22.4%) | 65 (19.9%) |  |  |
| **No** | 747 (60.9%) | 2169 (54.0%) | 1842 (57.1%) | 794 (67.8%) | 251 (76.8%) |  |  |

### Stress

High stress levels were observed across all age groups (Fig. 1). Participants within the 18-24 and 25-34 age groups exhibited comparable levels of stress, as indicated by the PSS-10 questionnaire, suggesting a consistent stress response pattern among younger adult cohorts. Among these age cohorts, only 2.6% reported low stress levels, while 42.7-44.6% experienced moderate and 52.7-54.6% high levels of perceived stress (Fig. 1A). Advancing age is associated with a noticeable decrease in high perceived stress: 43.7% among persons aged 45-54 years and 38.5% among 55+ years old participants.

**Figure 1.**
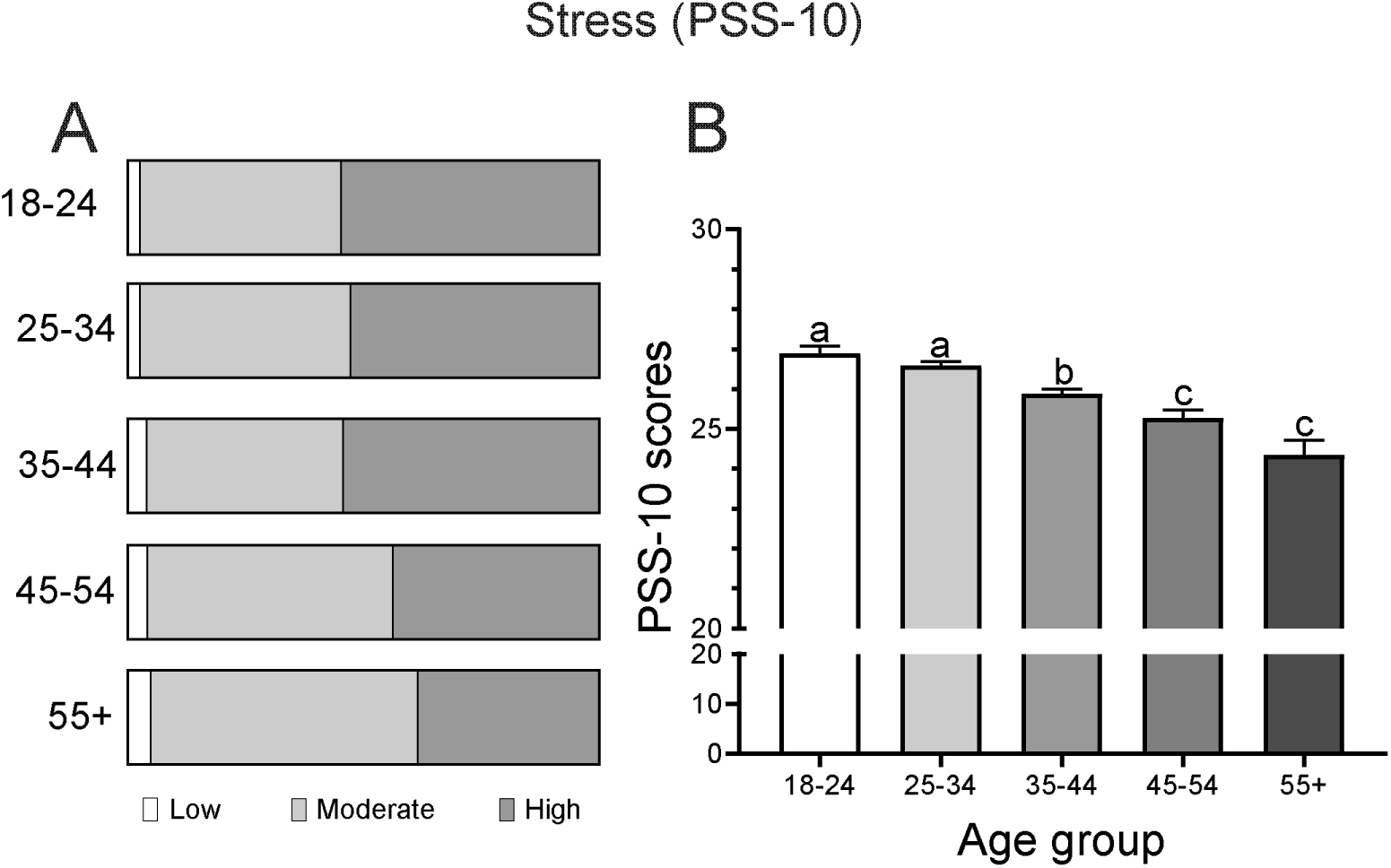
The prevalence of perceived stress among the respondents aged 18-24, 25-34, 35-44, 45-54, and 55+ years. (A) The proportion of individuals classified as experiencing low (0-13), moderate (14-26), or high (27-40) perceived stress according to the PSS-10 (Perceived Stress Scale). (B) PSS-10 scores across age groups. Statistically significant differences between groups (*p* < 0.05) were determined using the Kruskal-Wallis test followed by Dunn’s multiple comparison test. Distinct letters indicate significant pairwise differences.

Analysis of the total scores obtained from the PSS-10 questionnaire further confirmed differences between the groups (Fig. 1B). Persons of younger age groups including 18-24 and 25-34, exhibited higher stress levels (18-24 years: mean = 35.7, s.d. = 18.5; 25-34 years: mean = 34.3, s.d. = 17.8) compared to 35-44 group (mean = 32.7, s.d. = 17.6), while persons of older age groups (45-54 and 55+) showed the lowest levels of perceived stress (45-54 years: mean = 31.4, s.d. = 17.2; 55+ years: mean = 28.9, s.d. = 16.6).

### Anxiety

The findings of this study indicate a considerable prevalence of anxiety symptoms among the surveyed population, with notable differences observed between groups depending on their age. The assessment of anxiety symptoms across the study population revealed that moderate anxiety affected approximately 25.38 to 29.04% of individuals, while severe anxiety was reported by 13.76% to 20.06% of respondents (Fig. 2A). Statistically significant differences in anxiety severity were identified among the younger and more advanced groups (Fig. 2B). Participants aged 18-24 and 25-34 years demonstrated the highest mean anxiety scores (18-25: mean = 10.7, s.d. = 5.30; 25-34: mean = 10.5, s.d. = 5.15), whereas the lowest values were observed among 55+ persons (mean = 9.28, s.d. = 4.98). Individuals in the 45-54 age group exhibited low anxiety scores (mean = 9.66, s.d. = 5.20), with no significant difference when compared to the 55+ group. Based on the GAD-7 classification, between 39.1% and 49.1% of individuals across the groups exhibited moderate to severe anxiety, indicating the need for additional psychological evaluation and potential clinical follow-up.

**Figure 2.**
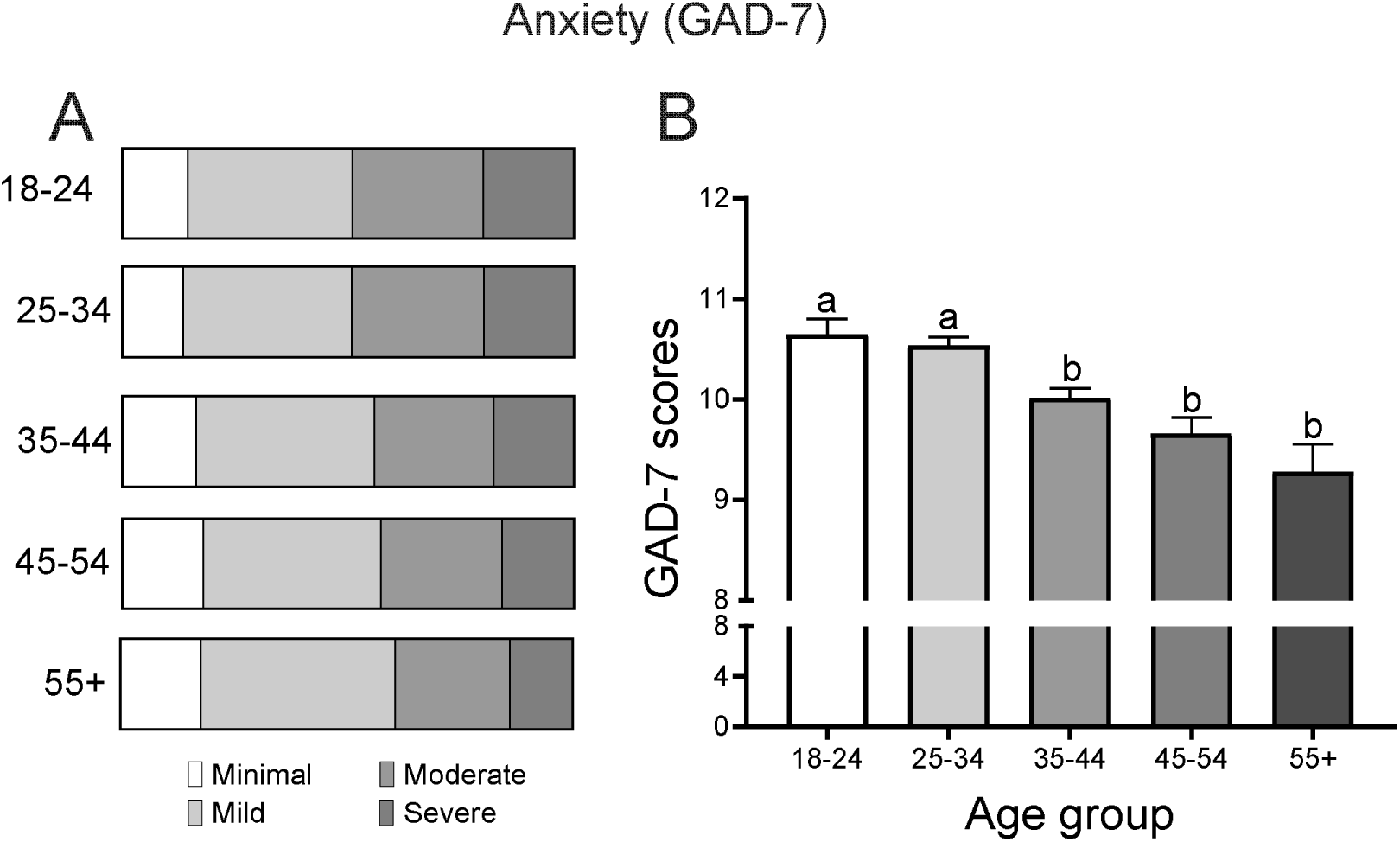
The prevalence of anxiety among the respondents aged 18-24, 25-34, 35-44, 45-54, and 55+ years. (A) The proportion of individuals with minimum (0-4), mild (5-9), moderate (10-14), or severe anxiety (15-21) according to the GAD-7 questionnaire. (B) Average GAD-7 scores across age groups. Statistically significant differences between groups (*p* < 0.05) were determined using the Kruskal-Wallis test followed by Dunn’s multiple comparison test. Distinct letters indicate significant pairwise differences.

### Depression

Assessment of the distribution of depression using the PHQ-9 revealed varying levels of depressive symptoms across the study groups. The proportion of individuals with moderate to severe depression ranged from 47.4 to 85.7%, indicating a substantial mental health burden (Fig. 3A). A gradual decline in depression severity was observed with increasing age. This pattern may be attributed to age-related differences in resilience, life experience, and cumulative adaptation to stress.

**Figure 3.**
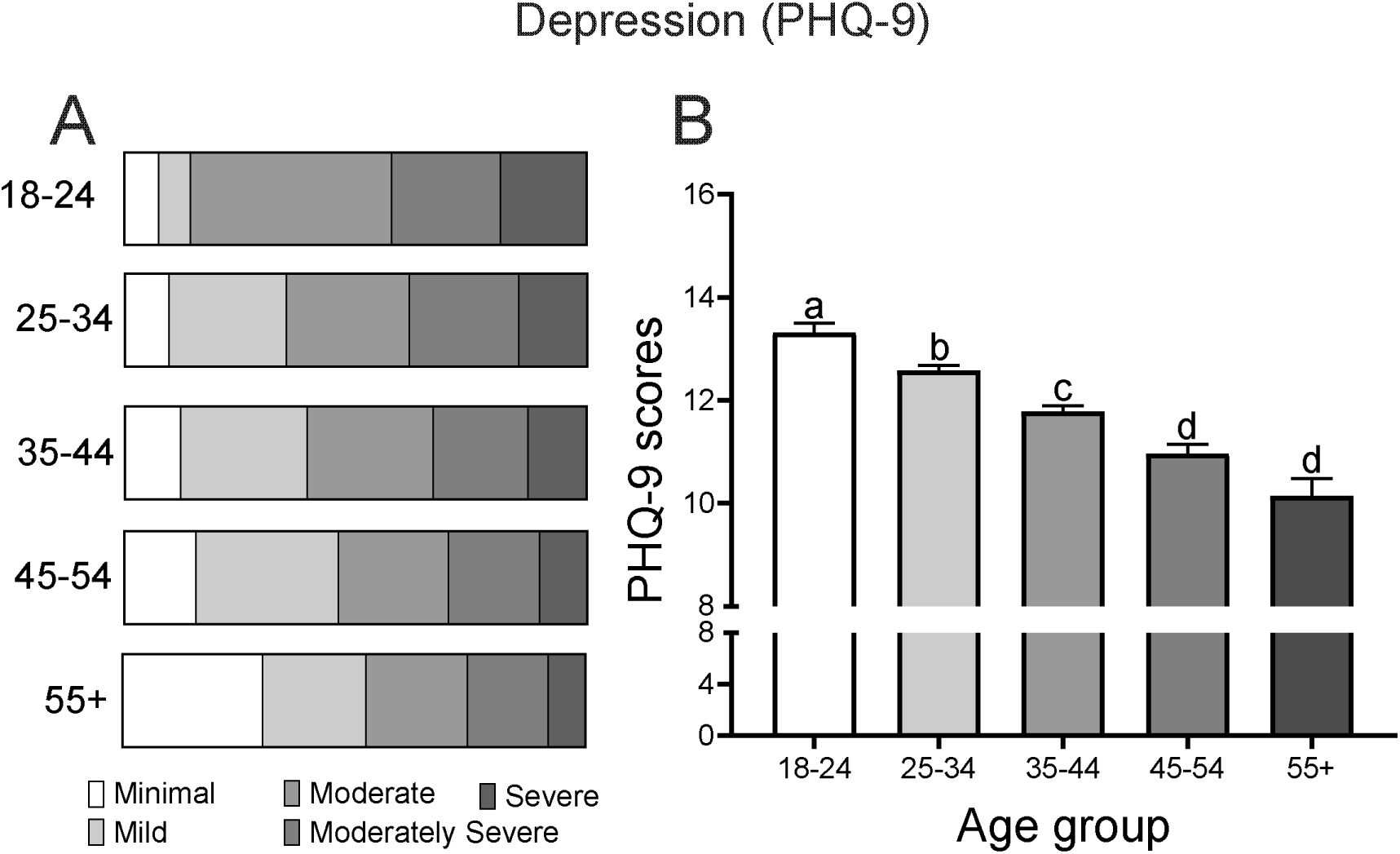
The prevalence of depression among the respondents aged 18-24, 25-34, 35-44, 45-54, and 55+ years. (A) The proportion of individuals with minimal (0-XX), mild (), moderate (), moderately severe () and severe depression () according to the PHQ-9 (Patient Depression Questionnaire). (B) PHQ-9 scores across age groups. Statistically significant differences between groups (*p* < 0.05) were determined using the Kruskal-Wallis test followed by Dunn’s multiple comparison test. Distinct letters indicate significant pairwise differences.

Group comparisons demonstrate statistically significant differences in depression severity, with the highest mean scores observed among participants of the 18-24 group (mean = 13.3, s.d. = 6.19) and the lowest among participants of the 55+ group (mean = 10.2, s.d. = 6.03) (Fig. 3B). Moreover, PHQ-9 scores decline progressively with age. The differences between each consecutive age group were statistically significant.

### Post-traumatic stress disorder

To assess PTSD prevalence, the PCL-5 questionnaire was applied. The results of the study revealed a substantial burden of post-traumatic stress symptoms across all groups, with the most severe manifestations observed among younger age individuals (18-24 and 25-34 years) (Fig. 4A). It was found that between 37.3 and 52.6% of participants met the criteria for probable PTSD (Fig. 4A). Based on the DSM-V criteria for PTSD diagnosis (Fig. 4B), we found that a substantial proportion of respondents, ranging from 38.8% to 52.5%, exhibited a level of symptoms consistent with clinical PTSD.

**Figure 4.**
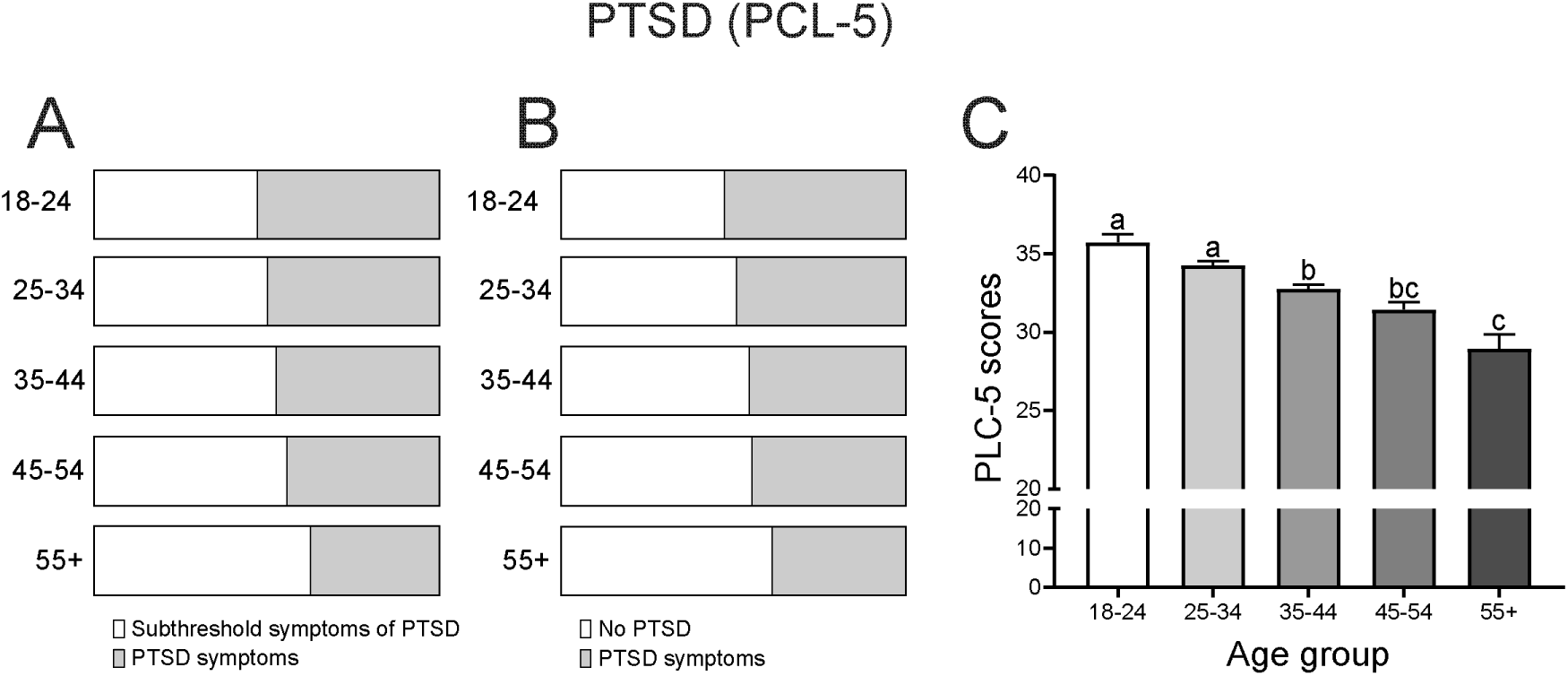
The prevalence of post-traumatic stress disorder among the respondents aged 18-24, 25-34, 35-44, 45-54, and 55+ years. (A) The proportion of individuals without (0-33) and severe PTSD (33-80) according to the PCL-5 (PTSD Check List) questionnaire. (B) Criteria-based classification of respondents: no PTSD and severe PTSD (per cent). (C) Average PCL-5 scores across age groups. Statistically significant differences between groups (*p* < 0.05) were determined using the Kruskal-Wallis test followed by Dunn’s multiple comparison test. Distinct letters indicate significant pairwise differences.

A gradual decline in PTSD prevalence was observed across age groups, with the highest rates among participants aged 18-24 and the lowest among those aged 55 and older (Fig. 4C). Comparative analysis indicated that individuals of 18-24 and 25-34 groups exhibited significantly higher average PCL-5 scores (18-24: mean = 35.7, s.d. = 18.5; 25-34: mean = 34.3, s.d. = 17.8) than 35-44 (mean = 32.7, s.d. = 17.6), while 55+ demonstrated significantly lower scores (mean = 28.9, s.d. = 16.6) (Fig. 4C). Persons aged 45-54 showed intermediate scores (mean = 31.4, s.d. = 17.2), with no significant difference when compared to the 35-44 and 55+ groups.

The PTSD assessment method, based on DSM-5 symptom cluster criteria, revealed that individual PCL-5 symptom clusters showed additional group-specific differences (Fig. 5). Individuals in the younger age groups (18-24 and 25-34 years) consistently scored highest across all domains. Intrusion symptoms were significantly more pronounced and equally prevalent among 18-24 group (mean = 8.49, s.d. = 5.36) as compared to the 35-44 (mean = 7.74, s.d. = 4.99), 45-54 (mean = 7.63, s.d. = 4.84) and 55+ groups (mean = 7.20, s.d. = 4.67) (Fig. 5A). Avoidance behaviors were more characteristic of 18-24 group (mean = 3.51, s.d. = 2.58) relative to 35-44 group (mean = 3.17, s.d. = 2.38), with no significant distinction between the other three groups (Fig. 5B). Negative alterations in cognition and mood demonstrated a progressive decrease from 18-24 to 55+ years (Fig. 5C). Hyperarousal symptoms were the most elevated in younger individuals including 18-24 (mean = 10.8, s.d. = 5.79) and 25-34 groups (mean = 10.6, s.d. = 5.58), showing a progressive deline to 55+ (mean = 8.55, s.d. = 5.05) with all group comparisons reaching statistical significance (Fig. 5D).

**Figure 5.**
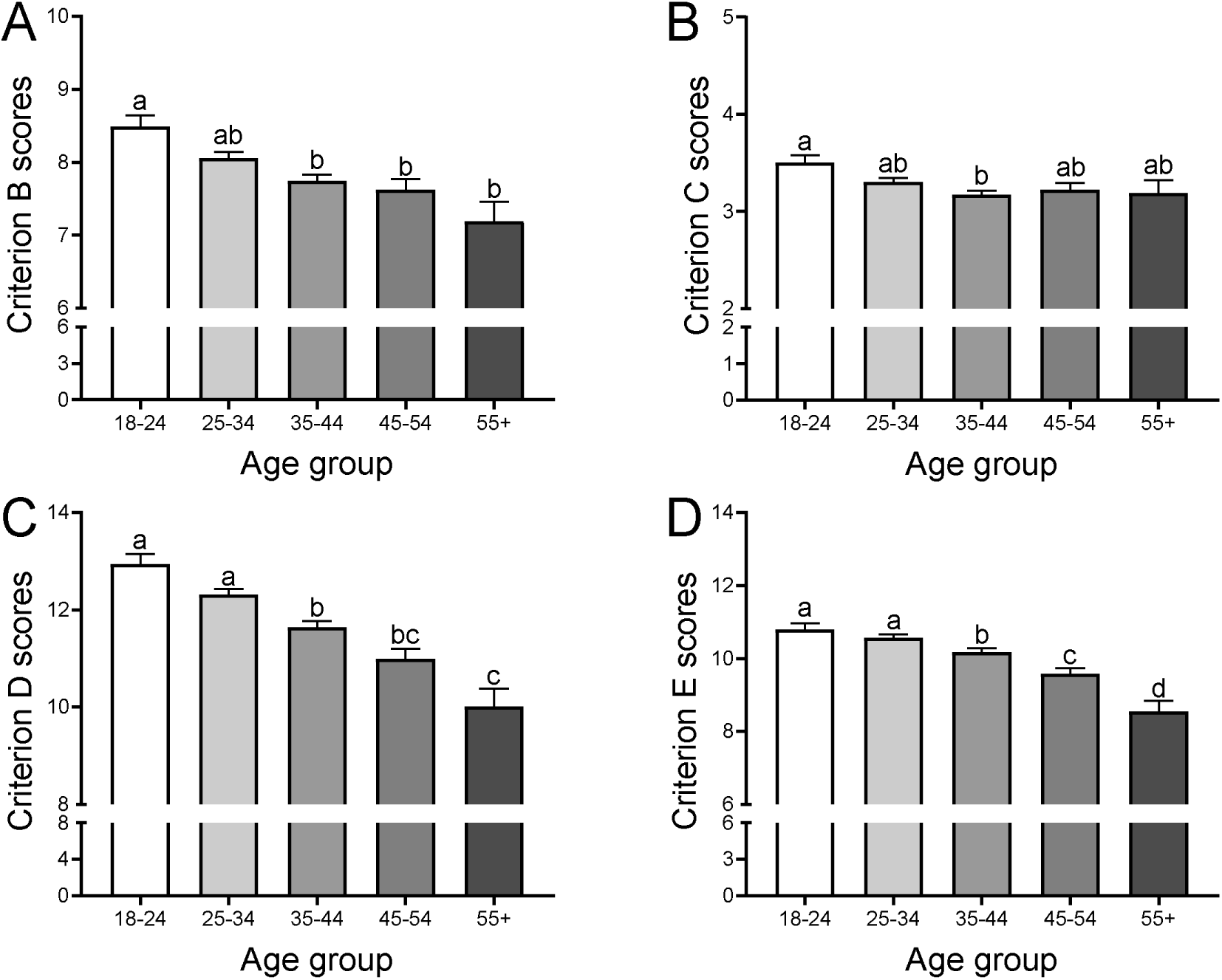
Comparative analysis of symptom cluster scores on the PCL-5 across study groups. (A) Intrusion symptoms (Criterion B), (B) Avoidance behaviours (Criterion C), (C) Negative changes in cognition and mood (Criterion D), and (D) Hyperarousal and reactivity (Criterion E). Statistically significant differences between groups (*p* < 0.05) were determined using the Kruskal-Wallis test followed by Dunn’s multiple comparison test. Distinct letters indicate significant pairwise differences.

## DISCUSSION

This study aimed to assess the mental health burden among adults in Ukraine two years after the full-scale Russian invasion in 2022. Symptom scores related to mental health disorders revealed that approximately 37.3-52.6% of Ukrainian adults met the threshold for probable post-traumatic stress disorder (PTSD), over 8% experienced severe depressive symptoms, and more than 17% reported severe anxiety symptoms. The present findings align with previous research indicating a substantial mental health burden among the Ukrainian population following the onset of conflict (Hyland et al., 2023; Karatzias et al., 2023; Khan & Altalbe, 2023; Kurapov et al., 2023; Lushchak et al., 2023). For instance, it was previously reported that about 40.5% of parents with children under 18 years exhibited probable PTSD or complex PTSD approximately six months after the invasion (Karatzias et al., 2023). We have previously reported on the prevalence of stress, anxiety, and PTSD symptoms among Ukrainian adults after 9-12 months of the 2022 Russian invasion of Ukraine (Lushchak et al., 2023). Our previous study found that 32.8-47.2% of group respondents had PTSD by the PCL-5 criterion (Lushchak et al., 2023). Our current results, collected two years post-invasion, suggest a further increase, with nearly half of the adult population meeting the criteria for probable PTSD.

Before the full-scale Russian invasion, estimates indicated that 8% of the general Ukrainian population met the criteria for PTSD (World Bank Group, 2017). Similarly, the pre-invasion prevalence rates for depression and anxiety were relatively low, at 6.3% and 3.2% respectively (World Bank Group, 2017), whereas our data reveal a marked increase in the severity and frequency of these symptoms among the adult population in the post-invasion context. Data collected within the first month of the Russian-Ukrainian war by Xu et al. (2023) indicated that 46.8% of respondents screened positive for depressive symptoms using the PHQ-2, while 54.1% showed elevated anxiety levels based on the GAD-2 assessment. Given the proximity to the traumatic events, these psychological reactions may represent immediate adaptive responses to extreme stress rather than long-term mental health disorders.

We also explored the distribution of the overall prevalence of key mental health disorders, including PTSD, depression, anxiety, and stress symptoms, across different age groups. We found that older adults (55+ years) exhibited lower levels of depressive, anxiety, and PTSD symptoms. At the same time, younger respondents consistently reported higher levels of psychological distress than their older counterparts. Our quantitative findings support these conclusions: the prevalence of severe depressive symptoms among participants aged 18-24 was 18.7% compared to 8% in those aged 55 and older, while severe anxiety prevalence similarly decreased from 20.0% to 13.8% across the same age groups. The findings suggest that younger adults, particularly those under 45, experience a greater mental health burden than middle-aged and even older individuals. This pattern aligns with findings from multiple recent studies conducted in conflict-affected populations, suggesting both developmental and contextual factors contribute to these differences.

It was previously reported that younger individuals often show greater emotional reactivity and distress following traumatic experiences (Stefaniak et al., 2021). The study of Kurapov and colleagues (2023) demonstrated that adult participants aged 28-45 showed the highest levels of anxiety, depression, PTSD, and CPTSD symptoms after six months of the Russian invasion. In contrast, younger Ukrainians (18-27 years old) and those aged 46-60 years reported higher resilience scores (Kurapov et al., 2023). An age of 28-45 is often associated with numerous personal and professional responsibilities, including caregiving, parenting, and career development. A combination of outside pressures associated with war and personal responsibilities can lead to higher levels of anxiety, depression, and trauma.

Previous epidemiological research has consistently demonstrated that the prevalence of PTSD tends to be lower among older adults compared to younger age groups (Kessler et al., 2005; Reynolds et al., 2016). For instance, data from large-scale population surveys in the United States show that lifetime PTSD rates are generally higher in younger individuals, with estimates around 6% in those aged 18-29, decreasing progressively to approximately 3% in individuals aged 60 and above (Kessler et al., 2005). Similarly, studies assessing 12-month PTSD prevalence also report a decline with advancing age, ranging from 4-5% in younger and middle-aged adults to about 3% among older adults (Reynolds et al., 2016). Data from the National Health and Resilience in Veterans Study (NHRVS) in the US revealed markedly elevated rates of probable lifetime PTSD among younger veterans, with an estimated 29.3% among those aged 18-29, decreasing to just 4% in veterans aged 60 and older (Wisco et al., 2016).

Biological and neurodevelopmental mechanisms may significantly contribute to the heightened vulnerability of younger adults to stress-related psychopathology. The prefrontal cortex (PFC), which plays a critical role in executive functioning, cognitive control, and emotional regulation, continues to undergo structural and functional maturation until near the age of 25 years (Casey et al., 2008). Neuroimaging studies have consistently shown that younger individuals exhibit greater amygdala activation in response to emotional or threatening stimuli (Wu et al., 2018) that may result in heightened emotional reactivity and impaired stress regulation (Wright et al., 2006). Furthermore, the hypothalamic-pituitary-adrenal (HPA) axis, which is involved in the physiological stress response, appears to be more reactive and better regulated in younger individuals (Kudielka et al., 2004). In situ hybridisation and autoradiography analysis across five human age groups revealed significantly higher levels of mRNA for glucocorticoid receptors (GR) in the prefrontal cortex during adolescence and young adulthood, and a decline in GR expression in older age groups (Perlman et al., 2007). This may suggest higher sensitivity to cortisol, the primary stress hormone, of younger adults as compared to older individuals.

Differences in coping resources, life experience, or the developmental stage of psychosocial identity formation may be among the contributing factors to higher mental health complications in younger individuals (Wood et al., 2018). Emotional maturity, accumulated life experience, and more established coping mechanisms may help older individuals manage stress more effectively. Younger adults often face additional challenges, especially during unstable conditions caused by war, such as disrupted education, unstable employment, and the interruption of key life transitions, which contribute to increased psychological strain. The uncertainty of the future and the burden of family responsibilities during ongoing conflict may contribute significantly to mental health complications in young participants. Additionally, younger individuals may have less developed coping resources and lower resilience, having had less life experience to process trauma and stress (Zapater-Fajarí et al., 2021). In contrast, previous life adversities or longer experience with stress management of older individuals may cause greater psychological resilience.

However, this age-related decline in symptom prevalence does not necessarily imply that older adults are unaffected. Previous research on war-affected populations has shown that older adults may experience delayed or somatised responses to trauma (Davison et al., 2006). Moreover, the response to traumatic factors in older individuals may differ from that of younger individuals and be manifested in a form of fatigue or social withdrawal. Our study also shows that individuals aged 55 and above demonstrated the highest proportion of non-displacement in the context of war-related migration, indicating a significant age-related pattern in displacement behaviour, potentially attributable to reduced mobility, established social ties, or increased place attachment within this demographic group.

This study provides the first empirical evidence of how repeated and prolonged exposure to armed conflict has affected mental health symptomatology among Ukrainians. We provide strong evidence that age is an important sociodemographic factor affecting individuals’ vulnerability to stress. Younger participants showed noticeably higher levels of depression, anxiety, and PTSD than older individuals, which may be due to differences in emotional regulation, life experiences, or coping strategies across age groups.

### Limitations

This study has several limitations related to both the characteristics of the sample and the overall research design. One notable limitation concerns the gender distribution of participants. Similar to other studies on the mental health of Ukrainians during wartime, our sample was predominantly female. Previous research in this area has also reported a high proportion of women respondents, ranging from 51 to over 80% (e.g., 51, 60, 82, and 83%) (Lushchak et al., 2023; Chudzicka-Czupała et al., 2022; Xu et al., 2022; Veldbrekht & Tavrovetska, 2022). While this trend may reflect greater willingness among women to participate in mental health research, it may also limit the generalizability of findings to the broader population, particularly men, whose psychological responses to conflict may differ. Moreover, gender-related differences may have influenced the study results, given recent evidence indicating that Ukrainian females in 2022 exhibited significantly higher levels of stress and anxiety compared to males (Chudzicka-Czupała et al., 2022).

## Contributors

OL and SB contributed to the conceptualisation of the study, the methodology, and the investigation. SB analyzed and validated the data. OL and OS contributed to data visualisation. OL, OS and KB prepared the original draft. All authors contributed to reviewing and editing. OL acquired funding, supervised the study and had full access to the raw data of the study. All authors have read and agreed to the published version of the manuscript.

## Funding

This work was partially supported by a grant from the Ministry of Education and Science of Ukraine [grant number 0123U101790].

## Data sharing statement

Access to original data can be obtained upon request to the corresponding author.

## Ethics committee approval

The study was approved by the Ethics Commission of the Psychological Department of Kryvyi Rih State Pedagogical University (protocol No. 12, 18.05.2023).

## Competing interests

The authors have no competing financial or non-financial interests to declare.

## ONLINE METHODS

### Methodology

An online survey was considered the most appropriate tool for this study to reach people across the country and abroad. The survey included gathering sociodemographic data (age, sex, education, current and prior location, social and mental health status) and four validated self-report questionnaires to measure stress, anxiety, depression and PTSD severities. All questionnaires had Ukrainian versions adapted and validated by earlier studies (Bezsheiko et al., 2016; Lushchak et al., 2023; Rzońca et al., 2024; Shevlin et al., 2018; Weigelt & Kizilhan, 2024).

The study was approved by the Institutional Ethics Committee of the Psychological Department of Kryvyi Rih State Pedagogical University (protocol No. 12, 18.05.2023).

### Procedure

The survey was distributed online through local and thematic groups on social media platforms, as well as through partnerships with bloggers and digital influencers, from October 12, 2023, to February 5, 2024. This timeframe covers a period of 20–24 months following the invasion. The distribution plan involved large groups of participants, asking them to fill out the survey form and inviting colleagues, friends, and relatives. Therefore, the initial invitation was sent to 50,000 people in total and included snowball sharing, with an unknown number of responses. Respondents clicked on the link to the questionnaire 22,231 times, and 11,014 surveys were completed in full. This resulted in a completion rate of 49.5%. The survey responses were collected by filling out a survey on Survio, a specialised platform for online surveys, along with written consent to participate in the study. Participation was voluntary and confidential.

### Stress

Stress was assessed using the Perceived Stress Scale (PSS-10), a ten-item questionnaire with scores ranging from 0 to 40 (Cohen, 1988). Respondents were asked to assess the frequency of the listed feelings and thoughts observed during the past month using the standard Likert scale from ‘never’ (0) to ‘very often’ (4). Obtained summed scores from 0 to 13 were considered as low stress, 14-26 as moderate, and 27-40 as high perceived stress (Veldbrekht & Tavrovetska, 2022). Cronbach’s alpha for PSS-10 was 0.867, and nd McDonald’s omega was 0.867.

### Anxiety

Anxiety was assessed using the GAD-7 questionnaire that estimates generalised anxiety disorder (Spitzer et al., 2006). This tool was validated for use in primary care and general populations (Williams, 2014) and has been utilised among Ukrainians as a reliable instrument, enabling data comparison with previous studies (Osokina et al., 2021; Lushchak et al., 2023). GAD-7 is a self-report seven-item anxiety questionnaire that determines the mental health state of an individual during the previous two weeks. The respondents assessed the degree of feeling nervous, anxious, unable to control worrying, easily irritable, and afraid that something might happen, ranking themselves as’not at all’, for’several days’, for’more than half the days’, or for’nearly every day’. Total scores from 0 to 21 were categorised as no or minimal anxiety (0-4), mild anxiety (5-10), moderate anxiety (11-15), and severe anxiety (16 and above). Cronbach’s alpha for GAD-7 was 0.891, and McDonald’s omega was 0.893.

### Depression

The severity of depressive symptoms was assessed using the Patient Health Questionnaire-9 (PHQ-9), a widely validated self-report instrument designed for screening and measuring the severity of depression (Kroenke et al., 2001). The PHQ-9 consists of nine items, each corresponding to one of the diagnostic criteria for major depressive disorder according to the DSM-IV. Participants rate the frequency of each symptom over the past two weeks on a 4-point Likert scale ranging from 0 to 3 (0 = “not at all”, 1 = “several days”, 2 = “more than half the days”, and 3 = “nearly every day”). The total score ranges from 0 to 27, with higher scores indicating greater severity of depressive symptoms. The score is interpreted as follows: 0-4 indicates minimal or no depression, 5-9 indicates mild depression, 10-14 indicates moderate depression, 15-19 indicates moderately severe depression, and 20-27 indicates severe depression.

### Post-traumatic stress disorder

Post-traumatic stress disorder was assessed using the PTSD Checklist for DSM-5 (PCL-5), developed for screening PTSD (VA.gov). PCL-5 is a self-assessment questionnaire with 20 items scored on a Likert scale from ‘not at all’ (0) to ‘extremely’ (4). The total PTSD symptom severity score, ranging from 0 to 80, was estimated by summing the scores of all questions. A cutoff of 33 was considered indicative of PTSD among samples, including Ukrainian adults (Bezsheiko et al., 2016). Another way to evaluate PTSD severity was based on separate criteria: A – trauma exposure or witnessing; B – intrusion symptoms; C – avoidance; D – negative alterations in cognition and mood; E – alterations in arousal and reactivity. The DSM-5 rule for PTSD diagnosis requires that at least one B item, one C item, two D items, and two E items should be rated as 3 (‘moderately’) or higher. Cronbach’s alpha for PCL-5 total scores was 0.941, and McDonald‘s omega was 0.941. Reliability of the other PCL-5 scales was also satisfactory (Criterion B – 0.870, McDonald’s omega – 0.871; Criterion C – 0.782, McDonald’s omega – 0.783; Criterion D – 0.883, McDonald’s omega – 0.884; Criterion E – 0.837, McDonald’s omega – 0.842).

## Data analysis

Data analysis was performed in SPSS Statistics 22 software (IBM, Armonk, NY, USA). The results were presented using descriptive statistics, Cronbach’s alpha coefficients for reliability calculation, the Shapiro-Wilk test to assess the normality of the distribution, the Kruskal-Wallis test for non-parametric data with further pairwise comparisons via the Mann-Whitney U-test, and the Pearson Chi-square test and Cramer’s V test for frequencies. Figures were generated in Prism GraphPad (GraphPad Software, MA, USA).

## Data Availability

All data produced in the present study are available upon reasonable request to the authors

## Notes

### Competing Interest Statement

The authors have declared no competing interest.

## References

Allwood MA, Bell-Dolan D, Husain SA. Children’s trauma and adjustment reactions to violent and nonviolent war experiences. J Am Acad Child Adolesc Psychiatry. 2002;41(4):450–7. doi: 10.1097/00004583-200204000-00018

Bezsheiko V. Adaptation of clinical-administered PTSD scale and PTSD Checklist for Ukrainian population. Psychos Med Gen Practice. 2016;1

Böttche M, Kuwert P, Knaevelsrud C. Post-traumatic stress disorder in older adults: an overview of characteristics and treatment approaches. Int J Geriatr Psychiatry. 2012;27(3):230–9. doi: 10.1002/gps.2725

Casey BJ, Jones RM, Hare TA. The adolescent brain. Ann NY Acad Sci. 2008;1124:111–126. doi: 10.1196/annals.1440.010.

Chudzicka-Czupała A, Hapon N, Chiang S-K, et al. Depression, anxiety and post-traumatic stress during the 2022 Russo-Ukrainian war, a comparison between populations in Poland, Ukraine, and Taiwan. Sci Rep. 2023;13:3602.

Cohen S. The social psychology of health. Sage Publications, Inc; Thousand Oaks, CA, USA: 1988. Perceived stress in a probability sample of the United States; pp. 31–67.

Davison EH, Pless AP, Gugliucci MR, King LA, King DW, Salgado DM, Spiro A, Bachrach P. Late-Life Emergence of Early-Life Trauma: The Phenomenon of Late-Onset Stress Symptomatology Among Aging Combat Veterans. Research on Aging. 2006;28(1):84–114. doi: 10.1177/0164027505281560

Ditlevsen DN, Elklit A. The combined effect of gender and age on post traumatic stress disorder: do men and women show differences in the lifespan distribution of the disorder? Ann Gen Psychiatry. 2010;9:32. doi: 10.1186/1744-859X-9-32.

El-Khodary B, Samara M, Askew C. Traumatic Events and PTSD Among Palestinian Children and Adolescents: The Effect of Demographic and Socioeconomic Factors. Front Psychiatry. 2020;11:4. doi: 10.3389/fpsyt.2020.00004

Frueh BC, Monnier J, Yim E, Grubaugh AL, Hamner MB, Knapp RG. A randomized trial of telepsychiatry for post-traumatic stress disorder. J Telemed Telecare. 2007;13(3):142–7. doi: 10.1258/135763307780677604

Harvard Medical School, 2007. National Comorbidity Survey (NCS). (2017, August 21). Retrieved from https://www.hcp.med.harvard.edu/ncs/index.php. Data Table 1: Lifetime prevalence DSM-IV/WMH-CIDI disorders by sex and cohort

Hyland P, Shevlin M, Brewin CR. The memory and identity theory of ICD-11 complex post-traumatic stress disorder. Psychol Rev. 2023;130(4):1044–1065. doi: 10.1037/rev0000418

Karatzias T, Shevlin M, Ben-Ezra M, McElroy E, Redican E, Vang ML, Cloitre M, Ho GWK, Lorberg B, Martsenkovskyi D, Hyland P. War exposure, post-traumatic stress disorder, and complex post-traumatic stress disorder among parents living in Ukraine during the Russian war. Acta Psychiatr Scand. 2023;147(3):276–285. doi: 10.1111/acps.13529

Kessler RC, Berglund P, Demler O, Jin R, Merikangas KR, Walters EE. Lifetime prevalence and age-of-onset distributions of DSM-IV disorders in the National Comorbidity Survey Replication. Arch Gen Psychiatry. 2005;62(6):593–602. doi: 10.1001/archpsyc.62.6.593

Khan AR, Altalbe A. Potential impacts of Russo-Ukraine conflict and its psychological consequences among Ukrainian adults: the post-COVID-19 era. Front Public Health. 2023;11:1280423. doi: 10.3389/fpubh.2023.1280423

Kroenke K, Spitzer RL, Williams JB. The PHQ-9: validity of a brief depression severity measure. J Gen Intern Med. 2001;16(9):606–13. doi: 10.1046/j.1525-1497.2001.016009606.x

Kudielka BM, Buske-Kirschbaum A, Hellhammer DH, Kirschbaum C. HPA axis responses to laboratory psychosocial stress in healthy elderly adults, younger adults, and children: impact of age and gender. Psychoneuroendocrinology. 2004;29(1):83–98. doi: 10.1016/s0306-4530(02)00146-4

Kurapov A, Kalaitzaki A, Keller V, Danyliuk I, Kowatsch T. The mental health impact of the ongoing Russian-Ukrainian war 6 months after the Russian invasion of Ukraine. Front Psychiatry. 2023;14:1134780. doi: 10.3389/fpsyt.2023.1134780

Lushchak O, Velykodna M, Bolman S, Strilbytska O, Berezovskyi V, Storey KB. Prevalence of stress, anxiety, and symptoms of post-traumatic stress disorder among Ukrainians after the first year of Russian invasion: a nationwide cross-sectional study. Lancet Reg Health Eur. 2023;36:100773. doi: 10.1016/j.lanepe.2023.100773

Maercker A, Forstmeier S, Wagner B, Glaesmer H, Brähler E. Posttraumatische Belastungsstörungen in Deutschland. Ergebnisse einer gesamtdeutschen epidemiologischen Untersuchung [Post-traumatic stress disorder in Germany. Results of a nationwide epidemiological study]. Nervenarzt. 2008;79(5):577–86. German. doi: 10.1007/s00115-008-2467-5.

Morina N, Stam K, Pollet TV, Priebe S. Prevalence of depression and post-traumatic stress disorder in adult civilian survivors of war who stay in war-afflicted regions. A systematic review and meta-analysis of epidemiological studies. J Affect Disord. 2018;239:328–338. doi: 10.1016/j.jad.2018.07.027

Osokina O, Silwal S, Bohdanova T, Hodes M, Sourander A, Skokauskas N. Impact of the Russian Invasion on Mental Health of Adolescents in Ukraine. J Am Acad Child Adolesc Psychiatry. 2023;62(3):335–343. doi: 10.1016/j.jaac.2022.07.845

Ozer EJ, Best SR, Lipsey TL, Weiss DS. Predictors of post-traumatic stress disorder and symptoms in adults: a meta-analysis. Psychol Bull. 2003;129(1):52–73. doi: 10.1037/0033-2909.129.1.52

Perlman WR, Webster MJ, Herman MM, Kleinman JE, Weickert CS. Age-related differences in glucocorticoid receptor mRNA levels in the human brain. Neurobiol Aging. 2007;28(3):447–58. doi: 10.1016/j.neurobiolaging.2006.01.010

Reynolds K, Pietrzak RH, Mackenzie CS, Chou KL, Sareen J. Post-Traumatic Stress Disorder Across the Adult Lifespan: Findings From a Nationally Representative Survey. Am J Geriatr Psychiatry. 2016;24(1):81–93. doi: 10.1016/j.jagp.2015.11.001.

Rzońca P, Podgórski M, Łazarewicz M, Gałązkowski R, Rzońca E, Detsyk O, Włodarczyk D. The prevalence and determinants of PTSD, anxiety, and depression in Ukrainian civilian physicians and paramedics in wartime-An observational cross-sectional study six months after outbreak. Psychiatry Res. 2024;334:115836. doi: 10.1016/j.psychres.2024.115836

Shevlin M, Hyland P, Vallières F, Bisson J, Makhashvili N, Javakhishvili J, Shpiker M, Roberts B. A comparison of DSM-5 and ICD-11 PTSD prevalence, comorbidity and disability: an analysis of the Ukrainian Internally Displaced Person’s Mental Health Survey. Acta Psychiatr Scand. 2018;137(2):138–147. doi: 10.1111/acps.12840

Spitzer RL, Kroenke K, Williams JB, Löwe B. A brief measure for assessing generalized anxiety disorder: the GAD-7. Arch Intern Med. 2006;166(10):1092–7. doi: 10.1001/archinte.166.10.1092

Steel Z, Chey T, Silove D, Marnane C, Bryant RA, van Ommeren M. Association of torture and other potentially traumatic events with mental health outcomes among populations exposed to mass conflict and displacement: a systematic review and meta-analysis. JAMA. 2009;302(5):537–49. doi: 10.1001/jama.2009.1132

Stefaniak AR, Blaxton JM, Bergeman CS. Age Differences in Types and Perceptions of Daily Stress. Int J Aging Hum Dev. 2022;94(2):215–233. doi: 10.1177/00914150211001588

VA.gov | veterans affairs. https://www.PTSD.va.gov/professional/assessment/adult-sr/PTSD-checklist.asp

Veldbrekht OO, Tavrovetska NI. Perceived Stress Scale (PSS-10): adaptation and approbation in the war circumstances. Problems Modern Psychol. 2022;2:16–27.

Viertiö S, Kiviruusu O, Piirtola M, Kaprio J, Korhonen T, Marttunen M, Suvisaari J. Factors contributing to psychological distress in the working population, with a special reference to gender difference. BMC Public Health. 2021 Mar 29;21(1):611. doi: 10.1186/s12889-021-10560-y. PMID: 33781240; PMCID: PMC8006634.

Weigelt A, Kizilhan JI. The Ukrainian version of the Perceived Injustice Questionnaire: A psychometric evaluation. Front Psychiatry. 2024;15:1446724. doi: 10.3389/fpsyt.2024.1446724

Williams N. The GAD-7 questionnaire. Occup Med. 2014;64:224. doi: 10.1093/occmed/kqu092

Wisco BE, Marx BP, Miller MW, Wolf EJ, Mota NP, Krystal JH, Southwick SM, Pietrzak RH. Probable Post-traumatic Stress Disorder in the US Veteran Population According to DSM-5: Results From the National Health and Resilience in Veterans Study. J Clin Psychiatry. 2016;77(11):1503–1510. doi: 10.4088/JCP.15m10188

World Bank Group, 2017. Mental health in transition: Assessment and Guidance for Strengthening Integration of Mental Health Into Primary Health Care and Community-Based Service Platforms in Ukraine. https://documents1.worldbank.org/curated/en/310711509516280173/pdf/120767-WP-Revised-WBGUkraineMentalHealthFINALwebvpdfnov.pdf (accessed October 5 2023).

Wood D, Crapnell T, Lau L, et al. Emerging Adulthood as a Critical Stage in the Life Course. 2017 November 21. In: Halfon N, Forrest CB, Lerner RM, et al., editors. Handbook of Life Course Health Development [Internet]. Cham (CH): Springer; 2018. Available from: https://www.ncbi.nlm.nih.gov/books/NBK543712/ doi: 10.1007/978-3-319-47143-3_7

World Health Organisation, WHO (2023b), Mental Health and Psychosocial Support (MHPSS) activities in countries hosting refugees from Ukraine: implementation of the international minimum standards for MHPSS, June.

Wright CI, Wedig MM, Williams D, Rauch SL, Albert MS. Novel fearful faces activate the amygdala in healthy young and elderly adults. Neurobiol Aging. 2006;27(2):361–74. doi: 10.1016/j.neurobiolaging.2005.01.014

Wu J, Geng X, Shao R, Wong NML, Tao J, Chen L, Chan CCH, Lee TMC. Neurodevelopmental changes in the relationship between stress perception and prefrontal-amygdala functional circuitry. Neuroimage Clin. 2018;20:267–274. doi: 10.1016/j.nicl.2018.07.022

Xu W, Pavlova I, Chen X, Petrytsa P, Graf-Vlachy L, Zhang SX. Mental health symptoms and coping strategies among Ukrainians during the Russia-Ukraine war in March 2022. Int J Soc Psychiatry. 2023;69:957–966.

Zapater-Fajarí M, Crespo-Sanmiguel I, Pulopulos MM, Hidalgo V, Salvador A. Resilience and Psychobiological Response to Stress in Older People: The Mediating Role of Coping Strategies. Front Aging Neurosci. 2021;13:632141. doi: 10.3389/fnagi.2021.632141

Zasiekina L, Zasiekin S, Kuperman V. Post-traumatic Stress Disorder and Moral Injury Among Ukrainian Civilians During the Ongoing War. J Community Health. 2023;48(5):784–792. doi: 10.1007/s10900-023-01225-5

